# Demographic Characteristics and Clinical Features of COVID-19 Patients Admitted in a Combined Military Hospital of Bangladesh

**DOI:** 10.1101/2022.02.12.22270893

**Authors:** Sabiha Mahboob, Fatema Johora, Asma Akter Abbasy, Fatiha Tasmin Jeenia, Mohammad Ali, Noor E Naharin, Jannatul Ferdoush

## Abstract

**Background:** COVID-19, one of the worst pandemics in humankind’s history on December 2019. Clinical presentations of COVID-19 patients are varied and being closely similar to those of seasonal flu, it’s difficult to differentiate it on first presentation as COVID. Clinical scenario and demographic characteristics provide important guideline in the management of COVID.

**Materials and Methods:** The objective of this cross-sectional study was to explore the demographic characteristics and clinical features of COVID-19 patients admitted in a Combined Military Hospital of Bangladesh. Data were collected from treatment records of patients of the CMH Bogura during the period of June 2020 to August 2020. Total 219 RT-PCR positive admitted patients were included as study population.

**Result:** Among 219 patients, 78.6% were male and 21.5% female. Mean age of patients was 34.3 ± 12.2. Highest percentages (67.2%) of patients were from age group 21-40 years. 85.4% of the patients had no comorbidities, and hypertension (10.1%) was the most common comorbidity. Most (83.1%) of the admitted patients were diagnosed as mild cases. 96.4% cases were symptomatic and fever (84.5%) was the most common symptoms of COVID, followed by dry cough (46.6%), sore throat (19.6%), headache (18.3%), bodyache (17.8%), loss of appetite (15.5%), tiredness (15.5%) and anorexia (14.2%).

**Conclusion:** This single center study revealed younger age, male predominance, less presence of comorbidites, mild cases, high proportion of symptomatic patients, and fever and cough as the most common presenting features among the admitted COVID-19 patients in CMH Bogura.

## INTRODUCTION

COVID-19, one of the worst pandemics in humankind’s history had hit us on December 2019. At first, few cases of atypical pneumonia were reported in Wuhan, the capital city of Hubei province of China, and later recognized as novel coronavirus infection caused by Severe Acute Respiratory Syndrome Coronavirus 2 (SARS-CoV-2).^1, 2^ World Health Organization (WHO) professed it as a global health emergency at first and later as pandemic on March 11, 2020. By September 2020, Total 220 coutries, areas or territories are affected with cases, more than 33 million are already infected and almost 100 thousands died because of COVID-19 pandemic. In Bangladesh, first case of COVID-19 was detected on 8 March 2020 and rapidly spreading out, total 360555 confirmed cases, including 5193 deaths.^3^ The severe acute respiratory coronavirus 2 (SARS-CoV-2) causing corona virus disease 2019 (COVID-19) pandemic is a foremost global health concern with a massive burden of disease worldwide.^4^

Knowledge of COVID-19 pathophysiology and transmission is changing as the pandemic evolves. SARS-CoV-2 is believed to spread primarily via respiratory droplets that are transmitted from person to person who are in close contact (usually within about 6 feet).^5^ The virus can also persist on surfaces to varying periods and degrees of infectivity. Asymptomatic persons account for approximately 40-45% of SARS-CoV-2 infection. These asymptomatic carriers can transmit infection to others for a long period, perhaps more than 14 days, and therefore, contribute to significant spread of the disease.^6^

The clinical scenario of COVID-19 is quite mixed, with the vast majority of patients being asymptomatic or only undergoing mild respiratory symptoms. The median incubation period of the disease is about four days ranging up to 14 days. Patients may present with constitutional symptoms (fever, headache, myalgia), upper respiratory tract symptoms (sore throat, rhinorrhea), lower respiratory tract symptoms (cough, dyspnea, chest tightness, sputum) and gastrointestinal symptoms (nausea, vomiting, diarrhea). The Covid-19 is very similar in symptomatology to other viral respiratory infections. Diagnosis of Covid-19 is commonly made through detection of SARS-CoV-2 RNA by PCR testing of a nasopharyngeal swab or other specimens, including saliva. Antigen tests are generally less sensitive than PCR tests but are less expensive and can be used at the point of care with rapid results.^5, 6, 7^ Cases vary from mild forms to severe ones that can lead to serious medical conditions or even death. About 5-15% of all patients with COVID-19 may progress to severe or critical illness, requiring sub-intensive or intensive care.^7^ The case fatality ratio varies across countries and depends on the health system, virulence of the strain, host immune response, genetic and environmental factors.^8^ The dissimilarity of demographic characteristics as well as clinical features vary between countries, and therefore different management strategy and treatment outcome has been observe.^9, 10, 11, 12, 13, 14, 15, 16^ There are few available data in this context from Bangladesh.^17, 18, 19, 20^ Hence, the present study was conducted to determine demographic characteristics and clinical features among the COVID-19 patients admitted in a combined military hospital of Bangladesh.

## MATERIALS &METHODS

This retrospective cross-sectional study was conducted in a Combined Military Hospital (CMH) Bogura of Bangladesh from June 2020 to August 2020. Ethical approval was taken from Institutional Review Board (IRB) of CMH Bogura. Data were collected from treatment records of COVID-19 patients of CMH Bogura. Investigators collected retrospective data from chronological register and treatment records kept in the Record Room of hospital. Data collection started from the most recently registered in-patient records to gradually backward manner. RT-PCR positive admitted COVID-19 patients of all age groups and sexes were included as study population. RP-PCR negative but radiologically suggestive cases of COVID-19 patients were excluded from the study. Treatment sheets of total 219 patients were included in this study. A review form was used for data collection. For maintenance of anonymity, an identification number was provided at top of the front page of the treatment sheet where confirmed clinical diagnosis and patient profile were mentioned. Data was compiled, presented and analyzed using Microsoft Excel 2007, and was expressed as percentage.

## RESULTS

**Table I** showing out of 219 patients, 78.6% were male and 21.5% were female. Mean age (years) of patients was 34.3 ± 12.2. Highest percentages (34.3%) of patients were from age group 21-30 years, followed by 31-40 years (32.9%).

**Table I.**
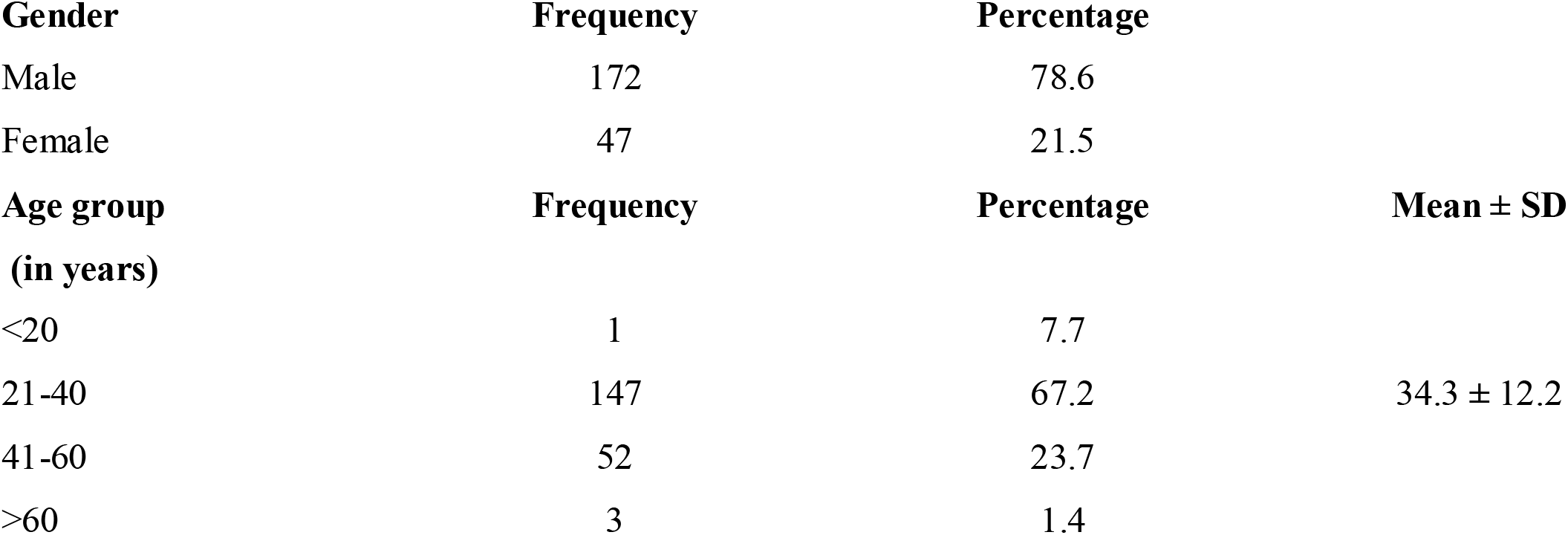
Demographic characteristics of admitted COVID-19 patients.

| Gender | Frequency | Percentage |  |
| --- | --- | --- | --- |
| Male | 172 | 78.6 |  |
| Female | 47 | 21.5 |  |
| Age group<br>(in years) | Frequency | Percentage | Mean ± SD |
| <20 | 1 | 7.7 |  |
| 21-40 | 147 | 67.2 | $34.3 \pm 12.2$ |
| 41-60 | 52 | 23.7 |  |
| >60 | 3 | 1.4 |  |

Figure I showing most (85.4%) of the patients has no comorbidity. Hypertension (10.1%) was the most common comorbidity, followed by diabetes mellitus (5.0%) and bronchial asthma (4.6%). **(Table II)**

**Figure 1.**
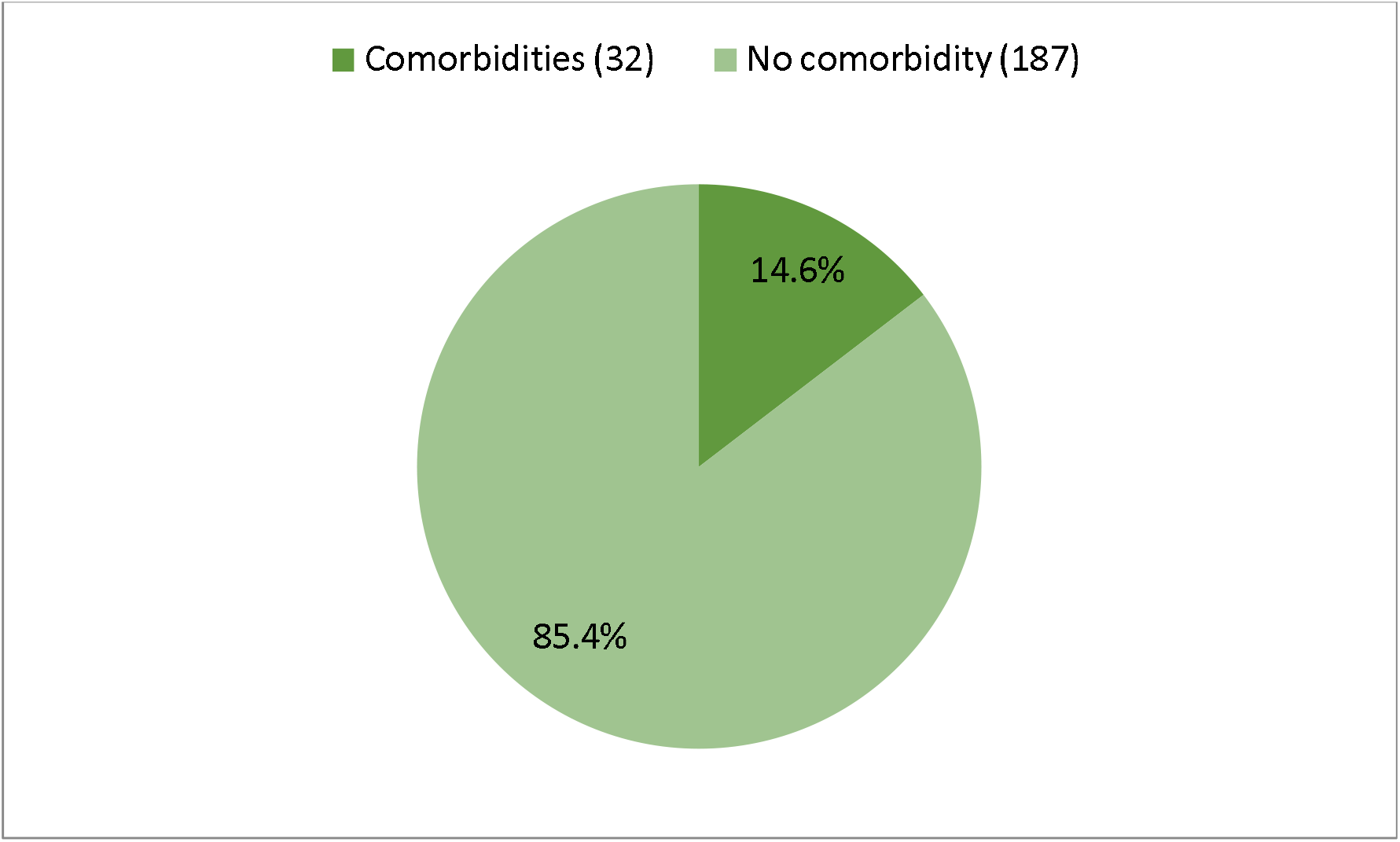
Proportion of patients having comorbidity.

**Table II.** Types of comorbidities.

| Number of comorbidities | Frequency | Percentage |
| --- | --- | --- |
| 1 | 18 | 8.2 |
| 2 | 7 | 3.2 |
| 3 or above | 7 | 3.2 |
| Specific co-morbidity | Frequency | Percentage |
| Hypertension | 22 | 10.1 |
| Diabetes mellitus | 11 | 5.0 |
| Bronchial asthma | 10 | 4.6 |
| Ischemic heart disease | 6 | 2.7 |
| Cerebrovascular disease | 2 | 0.9 |
| Hypothyroidism | 2 | 0.9 |
| Irritable bowel syndrome | 1 | 0.5 |
| Others | 2 | 0.9 |

Most (83.1%) of the admitted patients were diagnosed as mild cases (Figure 2). 96.4% cases were symptomatic (Figure 3).

**Figure 2:**
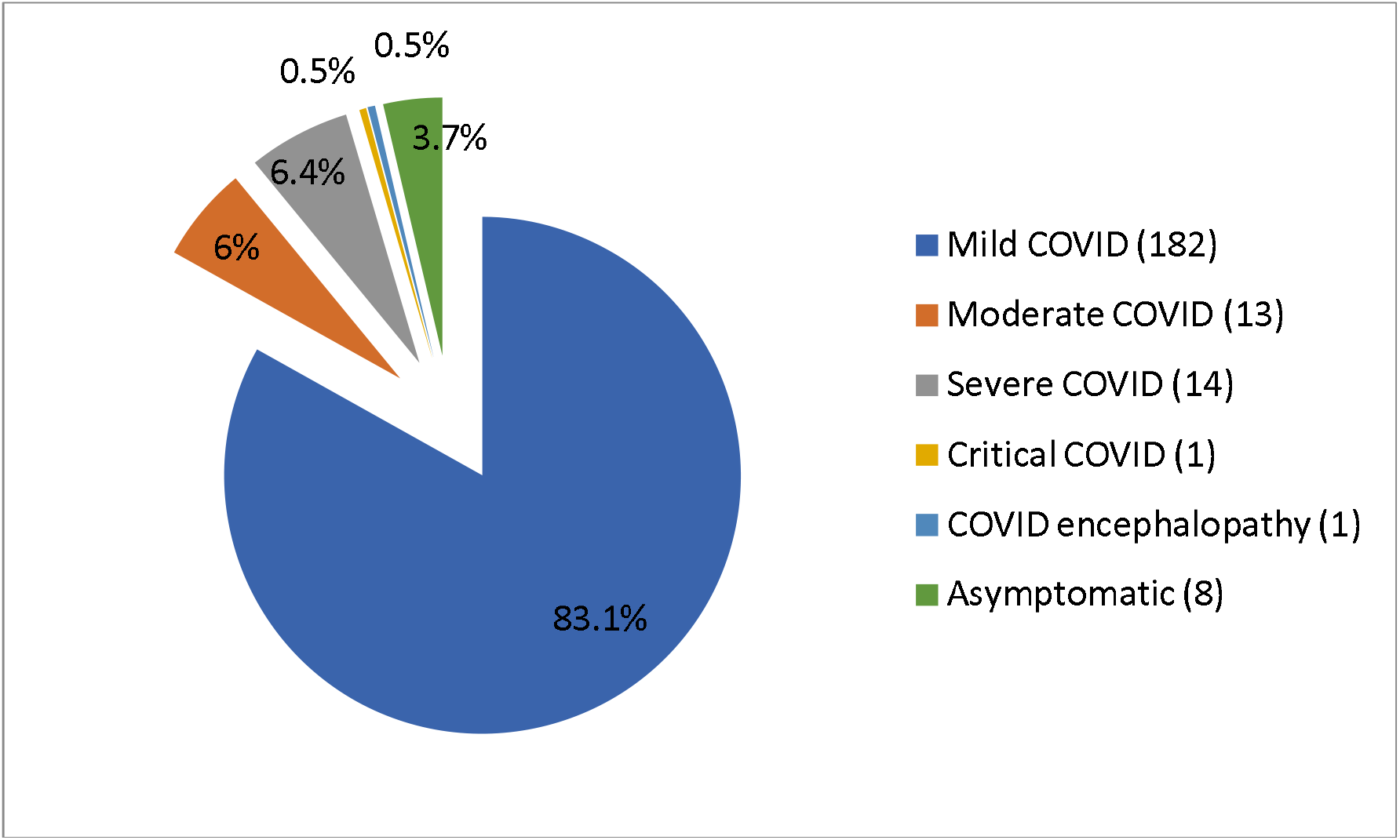
Diagnosis of COVID-19 patients.

**Figure 3:**
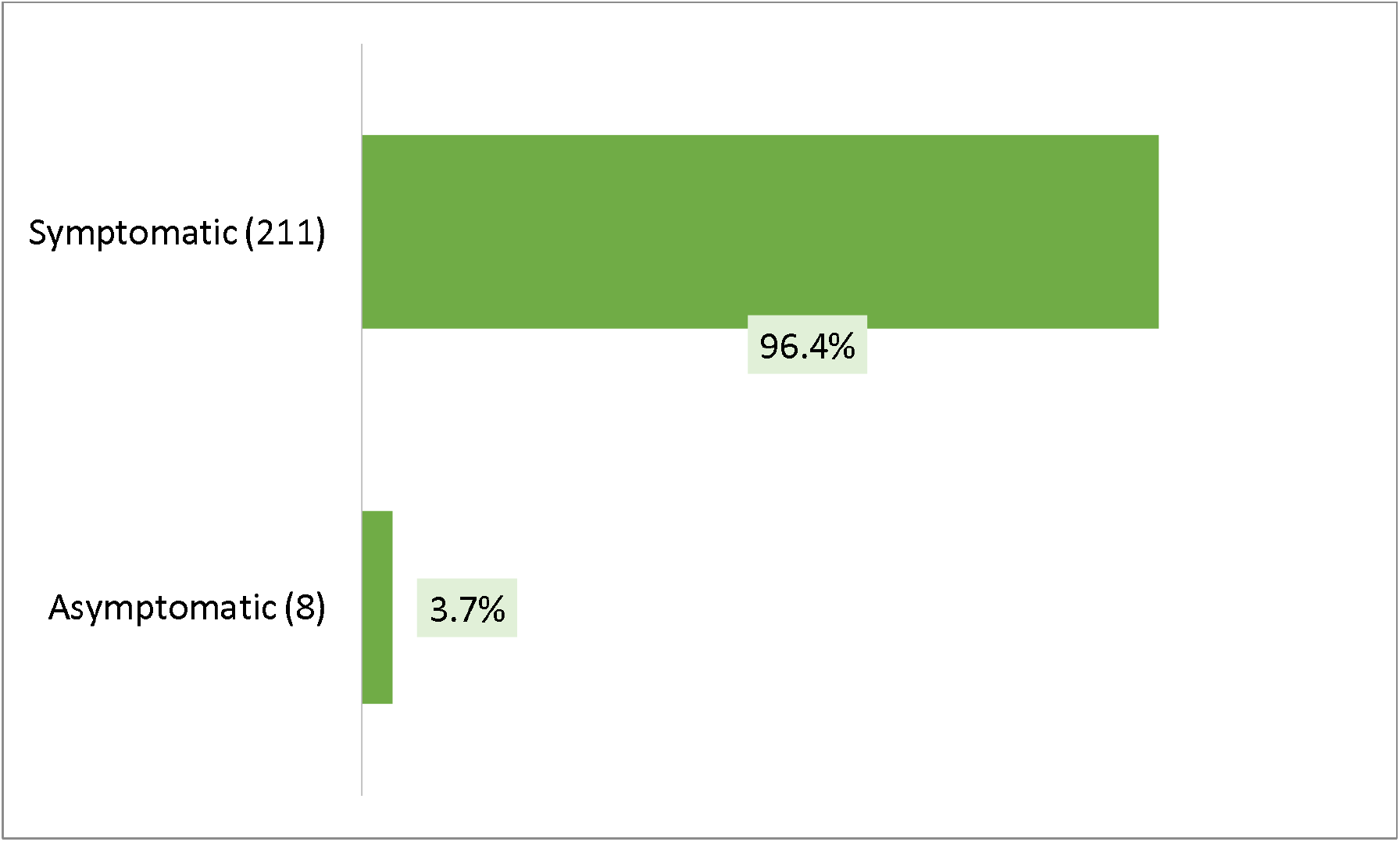
Proportion of asymptomatic and symptomatic cases of COVID-19.

**Table III** showed that fever (84.5%) was the most common symptoms of COVID, followed by dry cough (46.6%), sore throat (19.6%), headache (18.3%), bodyache (17.8%).

**Table III.** Presenting clinical features of COVID-19 patients on the day of admission.

| Variables | Frequency | Percentage |
| --- | --- | --- |
| Fever | 185 | 84.5 |
| Dry cough | 102 | 46.6 |
| Sore throat | 43 | 19.6 |
| Headache | 40 | 18.3 |
| Bodyache | 39 | 17.8 |
| Loss of appetite | 34 | 15.5 |
| Tiredness | 34 | 15.5 |
| Anorexia | 31 | 14.2 |
| Breathing difficulty | 28 | 12.8 |
| Loose motion | 21 | 9.6 |
| Nausea, vomiting | 13 | 5.9 |
| Loss of taste | 12 | 5.5 |
| Loss of smell | 7 | 3.1 |
| Runny nose | 11 | 5.0 |
| Chest pain | 4 | 1.8 |
| Insomnia | 2 | 0.9 |
| Vertigo | 1 | 0.5 |
| Restlessness | 1 | 0.5 |
| Irrelevant talk | 1 | 0.5 |
| Conjunctivitis | 1 | 0.5 |
| Abdominal pain | 1 | 0.5 |

## DISCUSSION

For last several months, world has been observing COVID-19 pandemic with enormous stress and anxiety. The clinical spectrum of SARS-CoV-2 infection appears to be wide, encompassing asymptomatic infection, mild upper respiratory tract illness and severe viral pneumonia with respiratory failure and even death, with many patients being hospitalized with pneumonia. The difference of socio-demographic characteristics between countries is also evident. The clinical presentation and outcomes of patients with COVID-19 are furthermore variable in different countries.^9, 10, 11, 12, 13, 19^ Therefore, the management strategy of one country needs to be individualized. During the initial phase of the COVID-19 outbreak, the diagnosis of the disease was complicated by the diversity in symptoms, imaging findings and in the severity of disease at the time of presentation. Therefore, this current study was conducted to identify the demographic characteristics and clinical features of COVID-19 patients admitted in a Combined Military Hospital of Bangladesh.

Current study found that most of the patients were in the age group of 21-40 (67.2%) and this finding was similar to two studies conducted in Bangladesh^19, 20^ but contrary to another study done in Bangladesh and also in China.^21, 13^ Mean age of study population was 34.3 ± 12.2 and that was concordance with studies conducted in Bangladesh and India.^22, 23^ But higher mean age was observed in most of the relevant researches.^17, 18, 19, 20, 21, 22, 24, 25^ and this was because current study was done in combined military hospital. In present study, male predominance (78.6%) was seen among COVID-patients. Similar male predominance was also found in several studies.^13, 18, 19, 20, 21, 22, 23, 24, 26^

Most of the patients (85.4%) of the current study had no comorbidities and 14.6% have one or more comorbidities. And that was similar to studies conducted by in Bangladesh and India.^19, 24^ Higher percentages of comorbid patients were observed in studies conducted by Hossain et al (66.5%), Revathishree et al (57%), Soni et al (29.8%) and Otuonye et al (49.4%).^21, 27, 23, 28^ Hypertension (10.1%) and diabetes mellitus (5.02%) were the most common comorbidities found in current research and that was concordance with relevant literatures.^19, 21, 22, 23, 24^ Severity of COVID-19 was categorized as asymptomatic, mild, moderate, severe and critical.^29^ Most of the patients of this study was diagnosed as mild case (83.1%) which mimic the findings of other studies.^9, 10, 13^, In current study, 3.7% of patients were asymptomatic but other studies found proportion of asymptomatic patients was ranging from 10.4% to 57.8%.^19, 23, 24, 28, 29, 30^ Among the symptomatic patients, the most frequent presentation was fever (84.5%), followed by dry cough (46.6%), sore throat (19.6%), headache (18.3%), body ache (17.8%), loss of appetite (15.5%), tiredness (15.5%) and anorexia (14.2%). These patterns of symptoms were closely similar to several studies done in home and abroad.^13, 19, 21, 22, 23, 28, 26, 31^

## CONCLUSION

Current study demonstrated demographic characteristics and clinical features of COVID-19 patients admitted in a combined military hospital in Bangladesh. The characteristic findings were younger age, male predominance, less presence of comorbidites, mild cases, high proportion of symptomatic patients, and fever and cough as the most common presenting features.

## Data Availability

All data produced in the present study are available upon request to the authors

## Notes

### Competing Interest Statement

The authors have declared no competing interest.

### Funding Statement

This study did not receive any funding

### Author Declarations

IRB of CMH Bogura, Bangladesh

